# Using Negative Control Outcomes to Detect Selection Bias in Mendelian Randomization Studies

**DOI:** 10.64898/2026.01.30.26345215

**Authors:** Apostolos Gkatzionis, George Davey Smith, Kate Tilling

## Abstract

Mendelian randomization is currently mainly implemented through the use of genetic variants as instrumental variables to investigate the causal effect of an exposure on an outcome of interest. Mendelian randomization studies are robust to confounding bias and reverse causation, but they remain susceptible to selection bias; for example, this can happen if the exposure or outcome are associated with selection into the study sample. Negative controls are sometimes used to detect biases (typically due to confounding) in observational studies. Here, we focus specifically on Mendelian randomization analyses and discuss under what conditions a variable can be used as a negative control outcome to detect selection mechanisms that could bias Mendelian randomization estimates. We show that the main requirement is that the negative control outcome relates to confounders of the exposure and outcome. Counter-intuitively, the effect of the negative control on selection is of secondary concern; for example, a variable that does not affect selection can be a valid negative control for an outcome that does. We also investigate under what conditions age and sex can be used as negative control outcomes in Mendelian randomization analyses. In a real-data application, we investigate the pairwise causal relationships between 19 traits, utilizing data from the UK Biobank. Treating biological sex as a negative control outcome, we identify selection bias in analyses involving commonly used traits such as alcohol consumption, body mass index and educational attainment.

## Introduction

Mendelian randomization (MR) uses genetic variants to explore the causal relationship between an exposure and an outcome of interest (Richmond and Davey Smith 2022, Sanderson et al. 2022). It is usually implemented as a form of instrumental variable analysis. MR has become an established approach in epidemiology since first formalized by Davey Smith and Ebrahim (2003). This has been facilitated by the availability of large-scale genetic databases, such as the UK Biobank, as well as the development of novel methods and accompanying software to aid researchers in conducting MR analyses (Yavorska and Burgess 2017, Hemani et al. 2018). As a result, the number of publications using MR has seen a rapid increase in recent years (Stender et al. 2024).

If all the instrumental variable (IV) assumptions are satisfied, MR analyses can overcome biases due to unobserved confounding and reverse causation that are prevalent in standard regression analyses. However, MR studies are still susceptible to other forms of bias. In particular, selection bias has been shown to affect instrumental variable and MR analyses, including in some situations when a multivariable regression analysis would not be affected (Canan et al. 2017, Gkatzionis and Burgess 2019, Hughes et al. 2019).

Selection bias exists when the value of a parameter of interest in the sample used for analysis differs from its value in the target population. When the parameter of interest is the exposure-outcome causal effect, there are two mechanisms by which selection bias may occur. The first is as a result of collider bias: two independent variables that are both causes of a third variable (the “collider”) will become artificially correlated in an analysis conditioned on a particular value of . This is illustrated in Figure 1. In practice, the collider may represent participation in a study or inclusion in the analysis sample, which motivates the use of collider bias as a structural framework for analysing selection bias (Hernan et al. 2004). A second type of selection bias occurs when the selection variable is not a collider, but the effect of interest is heterogeneous across levels of . This type of bias was first described by Greenland (1977) and has received increased attention in recent years (Hernan 2017, Lu et al. 2022, Mathur et al. 2025). Several mechanisms may give rise to this form of selection bias, including interaction, effect modification and non-collapsibility; here, we use the term “heterogeneity” to incorporate all of them. The impact of this form of bias on MR studies has received limited attention so far. It is worth noting that selection bias due to heterogeneity cannot occur under the sharp causal null hypothesis that the exposure does not affect the outcome for any individual in the target population (Hernan 2017); trivially, in this case, there would be no heterogeneity. These two types of selection bias are sometimes referred to as Type 1 and Type 2 selection bias respectively (Lu et al. 2022). In this paper, we will focus primarily on collider bias; as we discuss later, negative control outcomes are not suited to studying selection bias due to heterogeneity.

**Figure 1.**
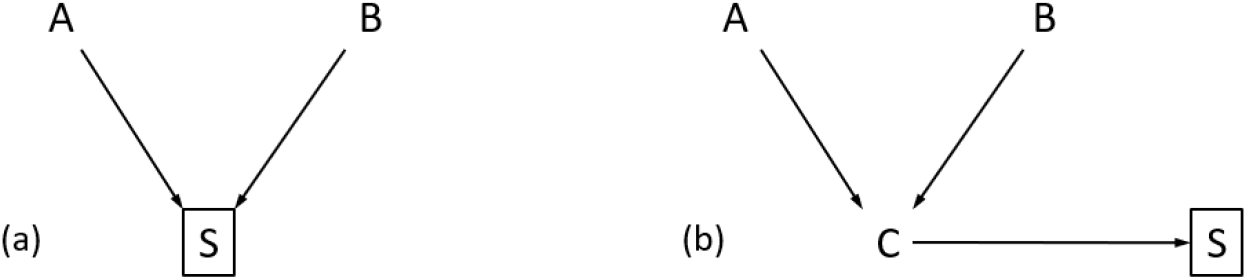
An illustration of collider bias. The variable S is a collider of A and B, as it is causally downstream of these two variables, and conditioning on S induces a spurious association between the parent variables A, B. This is the case both when A and B are “direct” causes of S (a) and when their effect on S is mediated by other variables (b).

Several large-scale genetic datasets, such as the UK Biobank (Fry et al. 2017, Schoeler et al. 2023), have been shown to suffer from non-random selection; MR studies utilizing those datasets could therefore be susceptible to selection bias of both types. This necessitates the development of diagnostics to detect and mitigate the bias.

In the field of epidemiology, it has long been proposed to use outcomes with expected null associations to uncover confounding and other forms of bias (Petitti and Kipp 1986, Davey Smith et al. 1992). In statistics, this approach was first discussed in terms of “known effects” (Rosenbaum 1989) but has more recently been labelled as the use of “negative controls” (Lipsitch et al. 2010, Davey Smith 2012). Intuitively, in a multivariable regression analysis estimating the association of an exposure with an outcome, a negative control outcome is a variable that is not caused by the exposure but for which the analysis is subject to the same biases as the analysis. Likewise, a negative control exposure should not be a cause of the outcome, but the analysis should suffer from the same biases as the original analysis. Several papers have provided guidance about the use of negative controls in regression analyses (Lipsitch et al. 2010, Arnold et al. 2016, Shi et al. 2020, Yang et al. 2024).

In the MR literature, negative controls are increasingly being used: a PubMed literature search using the terms “negative control” and “Mendelian randomization” returned 462 publications including both terms, more than half of which had been published within the last two years. Nevertheless, only a small number of publications have attempted to provide methodological advice regarding the use of negative controls in MR studies. Davies et al. (2017) proposed the use of negative control outcomes to compare and triangulate the results of MR analyses with those of conventional regression analyses. Motivated by advice given for regression analyses (Rosenbaum 1989), they suggested that a negative control outcome should be affected by the same confounders as the outcome of interest. Sanderson et al. (2021) explored the use of negative controls to detect bias due to population stratification. In that context, a suitable negative control outcome should be subject to the same influences from population structure as the MR outcome; in other words, confounders of the instrument and the outcome should also affect the negative control outcome. Both papers acknowledged that their results can be extended to MR analyses affected by selection bias but did not investigate this in detail. At the same time, Yang et al. (2021) discussed how to detect selection bias but focused on negative control exposures instead. More recently, Danieli et al. (2024) provided a rigorous theoretical framework for the use of negative control outcomes to detect violations of the instrumental variable assumptions, which encompasses different types of bias including collider bias. Building on these results, Dukes et al. (2025) investigated the use of negative controls to ensure identification of IV estimates under violations of the IV assumptions, while Orihara et al. (2024) developed a method that uses negative control outcomes to guide the selection of instruments for MR.

Some recent MR analyses have used age and sex as negative control outcomes. For example, this has been done as part of an ongoing debate around the performance of non-linear MR methods (Wade et al. 2023, Hamilton et al. 2024). Age and sex are appealing as negative control outcomes because they are commonly measured and likely to be available in most datasets, and it is easy to argue that they are not caused by the MR instruments, exposure and outcome. However, age and sex are also unlikely to be affected by other variables, including confounders of the exposure and outcome, therefore it may not be possible to replicate the biases of an MR analysis using age and sex as negative controls.

In this paper, we expand on the existing literature by considering the use of negative control outcomes to detect selection bias in instrumental variable and MR studies. Our results are presented in the context of MR studies using genetic instruments, but it should be noted that these results are also applicable to instrumental variable analyses using non-genetic instruments. Our work complements that of Sanderson et al. (2021) by focusing on selection bias instead of population stratification, and also complements that of Yang et al. (2021) by considering negative control outcomes instead of exposures. First, we illustrate how selection bias can affect MR studies using causal diagrams. We then consider for illustration a scenario where the MR exposure affects study participation and discuss what conditions a negative control outcome should satisfy in order to detect selection bias in the original MR analysis. We then generalize our findings to analyses where any combination of the instrument, exposure, confounder and outcome may affect study participation. We also discuss the use of age and sex as negative controls; we argue that, age and sex can be used to detect selection bias in some scenarios, but careful consideration should be given to their relationship with other analysis variables. Finally, we illustrate our results in a real-data application investigating selection bias in MR analyses between 19 commonly analysed biological traits, using data from the UK Biobank.

### Selection bias in MR studies

Suppose that our objective is to investigate the effect of an exposure *X* on an outcome *Y* in a target population of interest. MR uses a genetic instrument *G* to provide an estimate of the causal effect that is unaffected by unobserved confounders *U* of the exposure and outcome. For MR to yield an unbiased estimate of the *X* - *Y* causal effect, the instrument must satisfy the instrumental variable (IV) assumptions. Here, we use a version of the assumptions given by Pearl (2009): *G* is a valid instrument for the effect of exposure *X* on outcome *Y* in a directed acyclic graph (DAG) 𝒢if:

- (IV1): *G* is associated with *X* in DAG 𝒢.
- (IV2): *G* is conditionally independent of *Y* in the “manipulated DAG” derived from 𝒢 by eliminating the *X*-*Y* causal effect.

Figure 2 illustrates these assumptions. A third assumption is required to define conditional instruments, but will not be used in this paper; we report this in the Supplement and refer to (Pearl 2009) for more details. An additional fourth assumption of homogeneity or monotonicity is also necessary to assign a causal interpretation to point estimates obtained by an MR analysis, though that assumption is not required for testing the causal null hypothesis (Sheehan and Didelez 2020, Hartwig et al. 2023, Hernan and Robins 2024).

**Figure 2.**
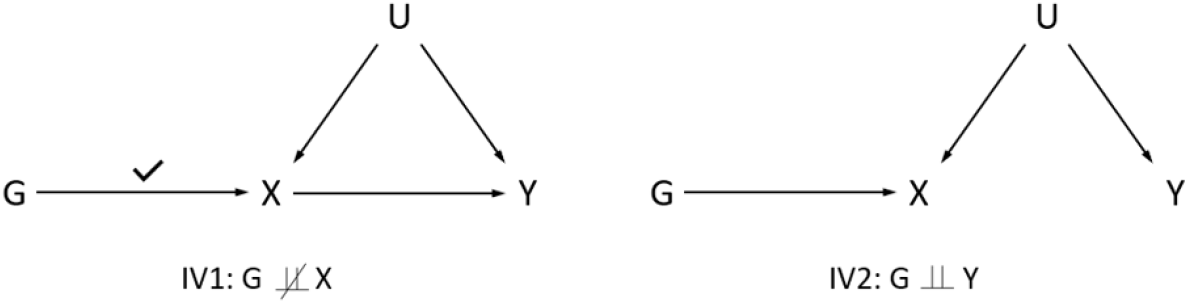
An illustration of Pearl’s IV assumptions. Left: the typical DAG used to represent an MR analysis. Assumption IV1 states that the instrument *G* must be associated with the exposure *X* in this DAG. Right: The “manipulated DAG”, derived by removing the exposure-outcome arrow. Assumption IV2 states that in this DAG, the instrument and the outcome are independent (i.e. there can be no open path connecting *G* and *Y*).

We note that several different ways of stating the IV assumptions have been used in the literature (Hernan and Robins 2006, Pearl 2009, Sanderson et al. 2022). The version we use here is relatively simple and flexible in detecting biases that can be represented in causal diagrams: in a DAG representing an MR analysis, bias will occur if there is an open path between the instrument and the outcome that does not include the “*X* → *Y*” arrow. We refer to the Supplement for some alternative versions of the IV assumptions and their usefulness for studying selection bias.

Several authors have investigated the impact of selection bias on MR analyses (Canan et al. 2017, Munafo et al. 2018, Gkatzionis and Burgess 2019, Hughes et al. 2019). These papers have focused on collider bias and illustrated that restricting an MR analysis to a selected sample (i.e. conditioning on selection/participation) can induce a non-causal association between the instrument and the outcome, in violation of assumption IV2. Consider, for example, the three causal diagrams of Figure 3. For simplicity, we will refer to one-sample MR analyses here; the theory presented in this paper applies to both one-sample and two-sample MR, but it is intuitively easier to think of “selection into the study sample” in the one-sample setting. Let *S* be a binary variable that represents selection: for every individual *i* in the target population, that individual is included in the study sample if *S*_*i*_ = 1 and not included if *S*_*i*_ = 0. In the DAG of Figure 3a, the MR exposure affects selection into the study. In such a study, the instrument and the confounder are both causally upstream of the selection indicator via their effects on the exposure. Running the MR analysis in the selected sample is equivalent to conditioning on . Therefore, even if is a valid instrument at a population level, it will become invalid in the selected sample because conditioning on opens the path between the instrument and the outcome and violates assumption IV2.

**Figure 3.**
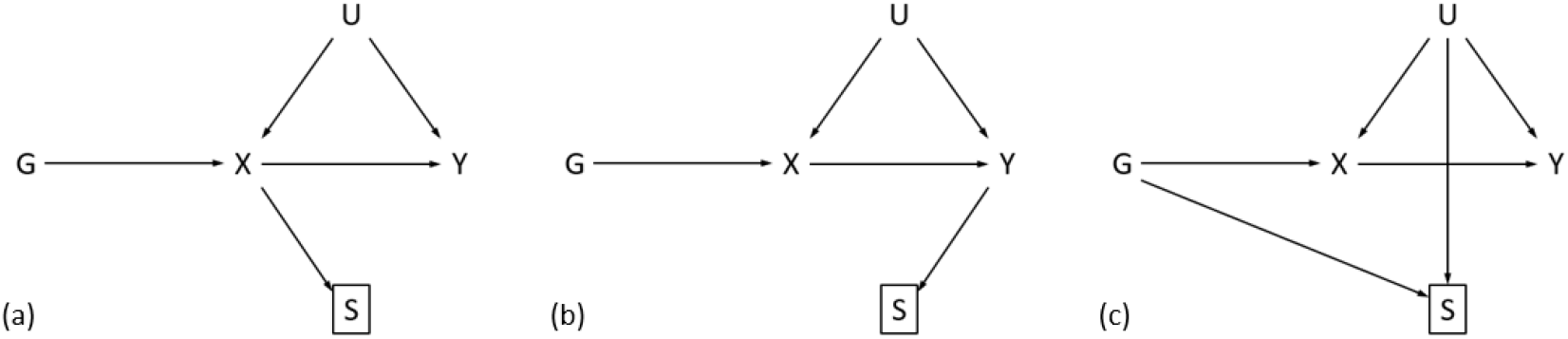
Causal diagrams representing MR analyses where selection (represented by the variable ) can induce collider bias. This can happen when (a) the exposure affects selection, (b) the outcome affects selection, or (c) the instrument and confounder independently affect selection.

The same can happen if the outcome of the MR study affects study participation (Figure 3b). In this case, the instrument is causally upstream of study participation via the path . Note, however, that this path is blocked if the MR exposure does not have a causal effect on the outcome, meaning that selection bias does not occur under the causal null hypothesis (Gkatzionis and Burgess 2019). Selection bias can also occur if the instrument and the confounder affect participation through different mechanisms and not via their effects on the exposure and outcome (Figure 3c). Finally, any combination of the above scenarios can also induce bias: for example, if study participation is influenced by both the exposure and the outcome, or by the exposure and the confounder, etc (Hughes et al. 2019)

The three DAGs of Figure 3 represent instances of collider bias: the bias is induced because selection is causally downstream of the instrument and confounder . However, as we have argued, selection bias can also occur due to heterogeneity. Although it is not possible to formalize this using DAGs, consider the diagram of Figure 4 as an example. Here, the exposure and the variable both affect the outcome . It is therefore possible that could act as an effect modifier for the effect of on ; we visualize this informally by a dotted arrow from to the causal effect. In addition, the effect modifier also affects the selection indicator . This setting is sometimes called effect modification by proxy (VanderWeele and Robins 2007). In this case, the unconditional causal effect will differ from the conditional effect given *S* = 1, and an IV analysis conducted in the selected sample will yield a biased estimate of the true *X*-*Y* causal effect. Intuitively, the causal effect is averaged across the distribution of *M*; but because *M* affects selection, that distribution differs between the target population and the selected sample. Bias can also be induced due to heterogeneity in the instrument-exposure relationship, although in this case the bias in the resulting MR estimate only occurs if the exposure has a causal effect on the outcome.

**Figure 4.**
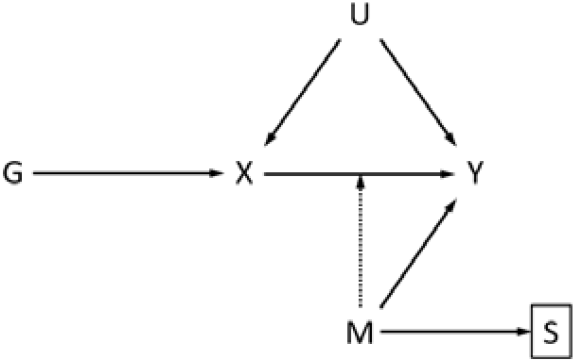
Diagram representing an MR analysis affected by selection bias due to heterogeneity. This happens because the variable acts as an effect modifier of the exposure-outcome causal effect (as represented by the dotted arrow) and also relates to selection .

### Negative Control Outcomes for Selection Bias

Here, we explore under what conditions a variable can be used as a negative control outcome to detect selection bias in MR studies. Similar to Sanderson et al. (2021), our objective is simply to detect the presence of bias and not to estimate its magnitude.

As discussed, our objective is to estimate the exposure-outcome causal effect in a target population of interest. We assume that *G* is a valid instrument for the (unconditional) MR analysis of exposure *X* on outcome *Y* in that population. However, for our analysis, we only have access to data from a (possibly non-representative) sample from that population. The binary indicator *S* indexes inclusion into the study sample. In this setting, a negative control outcome *N*_*Y*_ should satisfy two conditions:

i. *N*_*Y*_ should not be affected by the exposure. In other words, an MR analysis of *X* on *N*_*Y*_ in the target population should yield a null result.
ii. The MR analysis of *X* on *N*_*Y*_ should be affected by selection bias (and hence show evidence of an effect in the selected sample) if and only if the MR analysis of *X* on *Y* is affected by selection bias.

Here, we investigate how the negative control outcome *N*_*Y*_ may relate to the variables *G, X, U, Y, S* in order to satisfy these two conditions.

The first thing to note is that it is not possible to use negative control outcomes to detect selection bias due to heterogeneity. As we have argued, this form of bias does not occur under the (sharp) causal null hypothesis of no effect of the exposure on the outcome. However, a negative control analysis is by default conducted under the causal null hypothesis, as stated by condition (i) above. Therefore, negative control outcomes are only useful for studying collider bias, and that will be our focus in the rest of this manuscript.

For illustration we first consider the special case of Figure 3a, where selection is affected by the MR exposure, and then generalize our results to arbitrary selection mechanisms. As we have argued, an exposure-selection effect is enough to induce collider bias in the MR analysis. Our objective is to consider “candidate” negative control outcomes *N*_*Y*_ that relate to the variables *X, Y, G, U, S* in different ways and explore whether a negative control MR analysis using *X* as exposure and *N*_*Y*_ as outcome is also biased by selection.

Figure 5 illustrates various ways in which the negative control outcome may relate to . For each of these five variables, we consider three possible relationships: (a) causes that variable, (b) the variable and are confounded, or (c) that variable causes . This results in 15 different causal diagrams.

**Figure 5.**
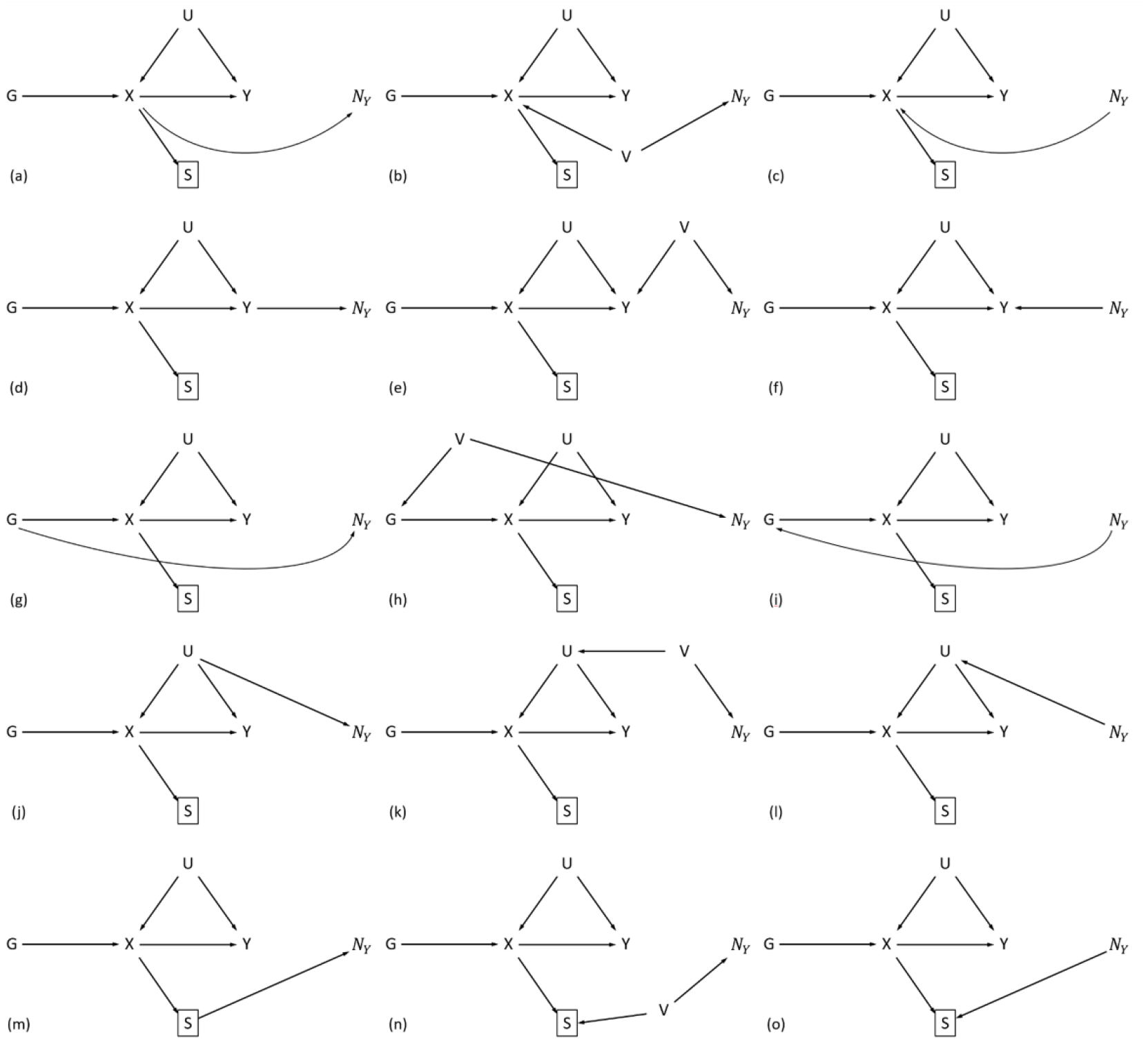
“Candidate” negative control outcomes for an MR analysis where the exposure affects selection into the study sample. The negative control outcome may relate to (top row of DAGs), (second row), (third row), (fourth row) or (bottom row). may be causally downstream (DAGs in the left column), confounded with (middle column) or causally upstream (right column) of these variables.

The causal diagrams of Figures 5a, 5d-5i and 5m represent cases in which would not be suitable for use as a negative control outcome. Specifically, in Figure 5a, the variable is causally downstream of the exposure. This means that the MR analysis would yield a non-null result at a population level, hence it invalidates as a negative control outcome. The same happens in Figure 5d, where is indirectly affected by the exposure via the path . In Figures 5e-5f, the variable either affects the original outcome or is confounded with it. This is not enough to induce bias in the conditional analysis as all paths from to collide in and is not a cause of the variable *S* that is conditioned on. In Figure 5g, it is the genetic instrument *G* that causes *N*_*Y*_; this induces bias due to pleiotropy in the *X*-*N*_*Y*_ MR analysis. However, this bias is not due to selection and can affect both a conditional (on *S)* and an unconditional MR analysis, therefore *N*_*Y*_ cannot be used to study collider bias in this case. Likewise in Figure 5h, where *N*_*Y*_ and *G* are confounded, or in Figure 5i, where *N*_*Y*_ acts as a cause of *G*; again, assumption IV2 is violated, but this violation is not due to collider bias. Finally, in Figure 5m, the selection indicator *S* affects *N*_*Y*_. This case is uncommon in practice as selection into a study is unlikely to affect other variables; if it were to happen, *X* would affect *N*_*Y*_ at a population level (indirectly via *S*) therefore *N*_*Y*_ would not be a valid negative control outcome.

The causal diagrams of Figure 5b-5c, 5j-5l and 5n-5o represent scenarios in which *N*_*Y*_ can be used to detect collider bias. In Figure 5b, the variable *N*_*Y*_ is confounded with the exposure *X* via the confounder *V*. An unconditional MR analysis of *X* on *N*_*Y*_ will be unaffected by the confounder and will show no evidence of an effect. However, an MR analysis conducted only in the selected sample will be biased because the path *G* → *X f*--*V* → *N*_*Y*_ becomes open conditional on *S*. In Figure 5c, the variable *N*_*Y*_ is a cause of *X*. Once again, this will not affect a population-level MR analysis of *X* on *N*_*Y*_ because MR is robust to reverse causation. However, the effects of *G* and *N*_*Y*_ on *X* collide, so conditioning on *S* opens a non-causal path from *G* to *N*_*Y*_ and induces bias. Figures 5j, 5k are similar to Figure 5b, as the negative control outcome is confounded with the MR exposure, while Figure 5l is similar to Figure 5c, as *N*_*Y*_ has a reverse causal effect on *X*. In all those cases, conditioning on *S* opens paths colliding in *X* and induces bias in the conditional analysis. In Figure 5n, *N*_*Y*_ relates to selection independently of *X*, via a common cause. In this case, the path *G* → *X* → *S f* ← *V* → *N*_*Y*_ becomes open in the selected sample, so bias is induced. Finally, in Figure 5o, collider bias occurs due to *N*_*Y*_ affecting selection.

To summarize, our exploratory analysis suggests two types of “candidate” negative control outcomes: variables that are causes of the MR exposure or share confounders with it, and variables that affect selection or share confounders with it. Combinations of these scenarios may also occur; for example, a “candidate” negative control outcome could share confounders with the MR exposure and also affect selection.

So far, we have focused on the scenario where the MR exposure affects selection into the study. In practice, negative control outcomes will be more useful for detecting collider bias in applications where it is not known which variables affect selection. Any combination of the instrument, exposure, confounder and outcome may affect selection into a study; this gives rise to 16 possible selection mechanisms. When the selection mechanism induces bias, the negative control outcome should allow us to replicate that bias in an exposure-negative control MR analysis. It is also possible that selection into the MR study happens in a way that does not induce bias in causal effect estimates; this could happen, for example, if the genetic instrument *G* affects selection, but the confounder, exposure and outcome do not. In this case, the negative control analysis should also be free of bias.

In Table 1, we list all 16 possible ways in which the variables *G, X, U* and *Y* could affect *S*. For each scenario, we first report whether the original MR analysis is subject to collider bias, by examining whether the IV assumptions are satisfied conditional on *S*. In total, collider bias occurs in 13 of the 16 scenarios.

**Table 1.** Bias in MR studies and the ability to detect it using seven “candidate” negative control outcomes. Boxes with bold font indicate scenarios where the negative control analysis yields a different result compared to the original MR analysis.

| Causes of $S$ | Bias in the $X - Y$ MR | Bias in the $X - N_Y$ MR analysis using outcome... | | | | | | |
| --- | --- | --- | --- | --- | --- | --- | --- | --- |
| | | $N_Y^1$ | $N_Y^2$ | $N_Y^3$ | $N_Y^4$ | $N_Y^5$ | $N_Y^6$ | $N_Y^7$ |
| None | No | No | No | No | No | No | No | No |
| G | No | No | No | No | No | No | Yes | Yes |
| X | Yes | Yes | Yes | Yes | Yes | Yes | Yes | Yes |
| U | No | No | No | No | No | No | No | No |
| Y | Yes | Yes | Yes | Yes | Yes | Yes | Yes | Yes |
| G, X | Yes | Yes | Yes | Yes | Yes | Yes | Yes | Yes |
| G, U | Yes | No | No | Yes | Yes | Yes | Yes | Yes |
| G, Y | Yes | Yes | Yes | Yes | Yes | Yes | Yes | Yes |
| U, X | Yes | Yes | Yes | Yes | Yes | Yes | Yes | Yes |
| U, Y | Yes | Yes | Yes | Yes | Yes | Yes | Yes | Yes |
| X, Y | Yes | Yes | Yes | Yes | Yes | Yes | Yes | Yes |
| G, U, X | Yes | Yes | Yes | Yes | Yes | Yes | Yes | Yes |
| G, U, Y | Yes | Yes | Yes | Yes | Yes | Yes | Yes | Yes |
| G, X, Y | Yes | Yes | Yes | Yes | Yes | Yes | Yes | Yes |
| U, X, Y | Yes | Yes | Yes | Yes | Yes | Yes | Yes | Yes |
| G, U, X, Y | Yes | Yes | Yes | Yes | Yes | Yes | Yes | Yes |

We then explore the suitability of seven candidate variables, 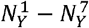 as negative control outcomes. These correspond to the causal diagrams of Figure 5 which were shown to yield valid negative control outcomes in the scenario where the exposure affects selection. Specifically:

- 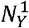 is confounded with the exposure *X* separately from *U* and does not affect *S* (Figure 5b).
- 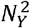 is a direct cause of *X* but does not affect selection *S* independently of *X* (Figure 5c).
- 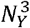 is confounded with *X* via *U* and does not affect *S* (Figure 5j).
- 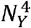 is confounded with *U* (hence also with *X*) and does not affect *S* (Figure 5k).
- 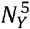 affects *U* and is therefore an indirect cause of *X* but does not affect *S* independently of *X* (Figure 5l).
- 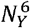 is confounded with *S* but is (unconditionally) unrelated to *X* (Figure 5n).
- 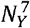 affects *S* but is (unconditionally) unrelated to *X* (Figure 5o).

For each of these variables, we explore whether collider bias is induced in an MR analysis using them as negative control outcomes, in each of the 16 selection mechanisms. We would like the negative control outcome analysis to be biased if and only if the original MR analysis is biased.

The results of Table 1 suggest that the variables 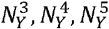, which are related to the confounder *U* of the exposure and outcome, are most suitable for use as negative control outcomes. Analyses using these variables exhibit the same pattern of bias as the original MR analysis, regardless of the selection mechanism. The other four “candidate” negative controls offer misleading results in at least one scenario. Variables 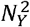 and 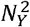 cannot detect collider bias when the instrument and the confounder cause selection (Figure 3c). These two variables are related to the exposure *X* but unrelated to the confounder *U*, therefore if bias is induced via *U*’s effect on *S*, they will not be able to detect it. On the other hand, variables 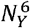, which shares a common cause with selection, and 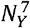, which is itself a cause of selection, falsely suggest the existence of bias in the scenario where the instrument causes selection.

Interestingly, our results imply that variables unrelated to selection into a study can be used as negative control outcomes to detect selection bias. The primary requirement is that the negative control should relate to confounders of the exposure and outcome.

It is worth noting that in two of the 16 scenarios of Table 1, the bias does not occur under the causal null hypothesis. These are the scenarios in which selection is affected by *Y* only, or by *U* and *Y*. In these two scenarios, the lack of an exposure-outcome causal effect will prevent bias both in the original MR analysis and in the negative control analysis.

It is also worth reiterating that the results of Table 1 relate only to collider bias, as this is the only form of selection bias that can be detected using negative control outcomes. For example, consider the scenario where only the confounder *U* affects selection into a study. Collider bias is not induced in this scenario. However, as *U* and *X* are common causes of the outcome *Y* It is possible that *U* may modify the *X* - *Y* causal effect, so selection bias can be induced due to heterogeneity. In the Supplement, we discuss this in more detail; we also explore how our work relates to graphical criteria for detecting selection bias in general (Daniel et al. 2012, Bareinboim et al. 2014, Mathur et al. 2025). Finally, in the Supplement, we briefly discuss selection bias in scenarios where the MR exposure and outcome relate to selection via common causes, instead of directly causing it; collider bias is less likely in these scenarios but bias due to effect modification may still occur.

In applications, multiple confounders of *X* and *Y* may exist; causal diagrams sometimes represent these using a single “composite” confounder variable *U* for simplicity. In principle, *N*_*Y*_ can be used to detect collider bias if it is only affected by some of these confounders. In practice, it will likely be better to choose negative control variables that share as many of the common causes of *X* and *Y* as possible, and with confounding effects of similar strength. In this case, the magnitude of collider bias will be similar in the original MR and in the negative control analysis, so if bias is detected in the latter it will likely have a noticeable impact on the former. Finally, note that if selection bias is caused solely due to the effect of a confounder on selection (this can be the case e.g. in the “G, U” scenario of Table 1), the negative control outcome should relate specifically to the confounder that affects selection.

### Age and Sex as Negative Controls

Age and sex are routinely measured in most epidemiologic datasets and have long been used as ways of sanity checking conclusions from epidemiological studies (Stocks 1935, Greenwood 1948). More recently they have been used as negative control outcomes in MR analyses (Wade et al. 2023, Davey Smith and Ebrahim 2024, Hamilton et al. 2024). The main argument for their use is that age and sex are unlikely to be causally downstream of other variables and will therefore be unaffected by the exposure and outcome of the MR analysis. Unfortunately, the same argument suggests that age and sex are unlikely to share confounders with the exposure and outcome. Referring to our results in Table 1, this implies that age and sex are unlikely to function like variables 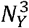 and 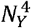, but may still affect confounders of the MR exposure and outcome and hence act like variable 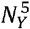 and therefore be suitable as negative control outcomes. Alternatively, age and sex may not relate to confounders but may affect the exposure, selection and/or outcome. In this case, they will function like variables 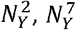 and Figure 5f respectively, or a combination thereof, and will not be valid as negative controls. Finally, in some applications it may be the case that age and sex do not relate to any of *G, X, U, Y, S*, and hence offer no information regarding bias in the MR analysis.

It is therefore possible for age and sex to be used as negative control outcomes for the purposes of detecting collider bias, but their use requires careful consideration of their relationship with the analysis variables instead of simply relying on the intuition that they are not causally downstream of the exposure and outcome.

Even if it is not possible to assess their relationship with confounding factors, age and sex may still be able to provide some information about the presence of collider bias, as it may be possible to assess their relationship with the exposure, outcome or selection. For example, information on the proportion of males and females in a particular sample will often be available; if these proportions differ from known proportions in the target population, one can argue that sex affects participation into the study sample. Combined with the fact that sex is not causally downstream of other variables, this will imply that sex is causally upstream of selection. By arguments similar to those used to construct Table 1, this will only yield misleading results in the scenario where the genetic instrument affects selection but the exposure, outcome and confounders do not. If that scenario can be excluded, based on subject matter knowledge or the availability of genetic data in unselected individuals, then a non-null effect in an MR analysis using sex as a negative control outcome can be taken as evidence of bias. Regardless, we suggest caution when using age and sex as negative controls. The use of causal diagrams can help researchers clarify what assumptions they are willing to make about their analyses and whether age and sex can be useful under these assumptions.

Finally, it is important to mention that even in scenarios where age and sex can provide information about collider bias, they may not be able to identify other forms of bias in the MR analysis, such as pleiotropy and population stratification. In practice, negative controls are often used agnostically to investigate biases of any form (Gutierrez et al. 2025), so other variables may be more suitable than age and sex.

### Real-data application: selection bias in MR analyses between 19 traits in UK Biobank

#### Data and methods

We conducted a real data analysis to illustrate our theoretical results about the use of negative controls for selection bias. Our analysis was based on data from the UK Biobank. Several authors (Fry et al. 2017, Batty et al. 2020, Schoeler et al. 2023, van Alten et al. 2024) have studied bias due to selective participation in UK Biobank. Schoeler et al. (2023) obtained data representative of the population of England from Health Survey England (HSE) and used them to compute weights for participation in UK Biobank. They then conducted weighted and unweighted GWAS for 19 commonly studied traits, using the UK Biobank data, and compared the results. They also conducted downstream MR analyses between pairs of traits and identified several exposure-outcome pairs for which the results of weighted and unweighted MR analyses differed, indicating selection bias. Finally, they investigated genetic associations with sex in their unweighted and weighted samples. Evidence of genetic effects on sex was detected in the unweighted GWAS, again indicating selection bias, but these effects attenuated in the weighted GWAS. Summary-level GWAS associations from that study are publicly available at https://drive.google.com/drive/folders/1SBJwsvlSSIOe_EpkY0HW0TFOVe6ywfxF.

Our real-data analysis was based on these summary data. First, we used sex as a negative control outcome and conduced MR analyses to estimate the effect of each of the 19 traits on sex, using the unweighted GWAS summary statistics. Then, we replicated the MR analyses of Schoeler et al. (2023) between each pair of traits, using both weighted and unweighted UK Biobank data, and identified pairs for which there was a clear difference in weighted and unweighted MR estimates, indicating selection bias. According to our theory, we would expect that traits affecting sex in the negative control analysis will show evidence of selection bias when used as exposures in the MR analyses.

The 19 traits investigated were BMI, height, alcohol consumption (number of drinks per week), smoking status, coffee intake, fruit intake, vegetable intake, education, physical activity, LDL cholesterol, systolic blood pressure, cancer, number of non-cancer diagnoses (“non-cancer”), diabetes, depression/anxiety, loneliness, reaction time, risk-taking and insomnia. For more details on how these traits were measured, we refer to Schoeler et al. (2023). For our MR analyses, we selected all SNPs associated with each of these 19 traits at the GWAS-significant level of 5x 10^-8^ in the corresponding unweighted GWAS. We then used those SNPs as instruments for both the weighted and the unweighted MR analyses. As in Schoeler et al. (2023), 6 of the 19 traits (vegetable intake, physical activity, cancer, depression/anxiety, loneliness, risk-taking) had fewer than 10 genetic variants passing the 5x 10^-8^ threshold in the corresponding GWAS, making it harder to instrument them effectively. Therefore, these six traits were not used as MR exposures and were only considered as outcomes.

#### Negative control analyses for sex

Table 2 contains MR estimates for the effect of each of the 13 exposures on sex, based on the unweighted and weighted GWAS data. The four exposures with the strongest evidence of association with sex in the negative control analysis (p < 0.05) were BMI, education, alcohol consumption and reaction time. The effect of alcohol consumption was strong enough to pass Bonferroni correction, while the other three effects were not. Of the four variables, education had the largest effect in absolute value. The effects of all four variables on sex attenuated in the weighted GWAS. There was weak evidence of a smoking-sex effect, but it did not pass the Bonferroni threshold for multiple testing.

**Table 2.** The effects of various traits on sex, using data from the unweighted and weighted GWAS of Schoeler et al. (2023).

| Exposure | Number of SNPs | Unweighted MR |  |  | Weighted MR |  |  |
| --- | --- | --- | --- | --- | --- | --- | --- |
|  |  | Estimate | 95% CI | P value | Estimate | 95% CI | P value |
| BMI | 227 | -0.002 | (-0.005, 0.000) | 0.033 | -0.002 | (-0.004, 0.001) | 0.202 |
| Height | 542 | 0.000 | (0.000, 0.001) | 0.219 | 0.000 | (-0.001, 0.001) | 0.594 |
| Alcohol | 21 | 0.005 | (0.002, 0.007) | 0.001 | 0.001 | (-0.002, 0.004) | 0.553 |
| Smoking | 40 | 0.017 | (-0.034, 0.069) | 0.511 | 0.055 | (0.005, 0.104) | 0.032 |
| Coffee | 19 | -0.001 | (-0.014, 0.013) | 0.902 | -0.008 | (-0.024, 0.008) | 0.346 |
| Fruit | 12 | 0.010 | (-0.016, 0.037) | 0.448 | 0.008 | (-0.040, 0.055) | 0.757 |
| Education | 218 | 0.009 | (0.002, 0.016) | 0.014 | 0.012 | (-0.001, 0.025) | 0.065 |
| LDL | 113 | 0.002 | (-0.007, 0.011) | 0.638 | 0.001 | (-0.013, 0.015) | 0.901 |
| SBP | 118 | -0.001 | (-0.001, 0.000) | 0.125 | -0.001 | (-0.002, 0.000) | 0.105 |
| Non-cancer | 14 | -0.025 | (-0.051, 0.002) | 0.067 | -0.020 | (-0.055, 0.015) | 0.264 |
| Diabetes | 37 | -0.068 | (-0.184, 0.049) | 0.256 | -0.103 | (-0.282, 0.077) | 0.262 |
| Reaction Time | 14 | -0.001 | (-0.001, 0.000) | 0.017 | 0.000 | (-0.001, 0.000) | 0.246 |
| Insomnia | 18 | 0.017 | (-0.061, 0.095) | 0.674 | 0.054 | (-0.053, 0.160) | 0.322 |

#### MR estimates

We implemented weighted and unweighted MR analyses for each of the 13 exposures with at least 10 genome-wide significant SNPs and each of the 19 outcomes, resulting in a total of 234 exposure-outcome pairs. We then compared the weighted and unweighted MR estimates for each pair. Schoeler et al. (2023) used a heuristic criterion to detect selection bias, according to which an absolute difference of more than 0.1 between (standardized) weighted and unweighted estimates was taken as indicative of bias. Here, we checked whether the 95% confidence intervals from the two MR analyses overlapped; for any pair of traits for which the confidence intervals did not overlap, we considered the unweighted MR estimate to be biased. This yielded four exposure-outcome pairs for which there was evidence of bias. These concerned the effects of education on smoking, depression, non-cancer diagnoses and insomnia. For all four traits, the bias acted in a positive direction, attenuating the risk-decreasing effects observed in the weighted MR towards zero. The direction of the effects remained unchanged.

For exploratory purposes, we also constructed 75% confidence intervals from the weighted and unweighted MR analyses and considered as “plausibly biased” any MR analyses in which these confidence intervals did not overlap. This yielded an additional seven exposure-outcome pairs: BMI-alcohol, BMI-smoking, BMI-education, height-physical activity, alcohol-vegetable intake, education-vegetable intake and education-reaction time. Estimates from the weighted and unweighted MR analyses for these exposure-outcome pairs are reported in Table 3. A full list of results for all 234 exposure-outcome pairs can be found in the Supplementary Data file.

**Table 3.** Exposure-outcome pairs for which we identified evidence of selection bias (i.e. the 95% or 75% confidence intervals from the weighted and unweighted MR analyses did not overlap).

| Exposure | Outcome | Number of SNPs | Unweighted MR |  |  |  | Weighted MR |  |  |  | Bias (95%) | Bias (75%) | Exposure-Sex P value |
| --- | --- | --- | --- | --- | --- | --- | --- | --- | --- | --- | --- | --- | --- |
|  |  |  | Estimate | P value | 95% CI | 75% CI | Estimate | P value | 95% CI | 75% CI |  |  |  |
| Education | Smoking | 218 | -0.087 | <0.001 | (-0.102, -0.073) | (-0.096, -0.079) | -0.151 | <0.001 | (-0.178, -0.124) | (-0.167, -0.135) | Y | Y | 0.014 |
| Education | Non-cancer | 218 | -0.178 | <0.001 | (-0.210, -0.147) | (-0.197, -0.160) | -0.285 | <0.001 | (-0.336, -0.233) | (-0.315, -0.254) | Y | Y | 0.014 |
| Education | Depression | 218 | -0.006 | 0.018 | (-0.010, -0.001) | (-0.008, -0.003) | -0.021 | <0.001 | (-0.030, -0.011) | (-0.026, -0.015) | Y | Y | 0.014 |
| Education | Insomnia | 218 | -0.056 | <0.001 | (-0.067, -0.044) | (-0.062, -0.049) | -0.090 | <0.001 | (-0.108, -0.072) | (-0.101, -0.080) | Y | Y | 0.014 |
| BMI | Alcohol | 227 | -0.070 | 0.034 | (-0.135, -0.005) | (-0.108, -0.032) | -0.156 | <0.001 | (-0.231, -0.081) | (-0.200, -0.112) | N | Y | 0.033 |
| BMI | Smoking | 227 | 0.022 | <0.001 | (0.018, 0.026) | (0.019, 0.024) | 0.015 | <0.001 | (0.010, 0.021) | (0.012, 0.019) | N | Y | 0.033 |
| BMI | Education | 227 | -0.090 | <0.001 | (-0.106, -0.075) | (-0.099, -0.082) | -0.063 | <0.001 | (-0.076, -0.051) | (-0.07, -0.056) | N | Y | 0.033 |
| Height | Physical activity | 542 | -0.005 | 0.003 | (-0.008, -0.002) | (-0.007, -0.003) | 0.001 | 0.685 | (-0.004, 0.006) | (-0.002, 0.004) | N | Y | 0.219 |
| Alcohol | Vegetable | 21 | -0.008 | 0.326 | (-0.023, 0.008) | (-0.017, 0.001) | 0.011 | 0.151 | (-0.004, 0.026) | (0.002, 0.020) | N | Y | 0.001 |
| Alcohol | Sex | 21 | 0.005 | 0.001 | (0.002, 0.007) | (0.003, 0.006) | 0.001 | 0.553 | (-0.002, 0.004) | (-0.001, 0.003) | N | Y | 0.001 |
| Education | Vegetable | 218 | -0.035 | 0.004 | (-0.059, -0.011) | (-0.050, -0.021) | 0.013 | 0.588 | (-0.033, 0.059) | (-0.014, 0.040) | N | Y | 0.014 |
| Education | Reaction time | 218 | -5.540 | <0.001 | (-7.482, -3.598) | (-6.680, -4.400) | -8.665 | <0.001 | (-11.822, -5.507) | (-10.518, -6.812) | N | Y | 0.014 |

#### Interpretation of results

All four MR analyses where there was a clear difference between the unweighted and weighted estimates used education as the exposure. Education was also one of the traits shown to be associated with sex in the unweighted GWAS, meaning that a sensitivity analysis using sex as a negative control outcome would be able to identify the bias in unweighted MR estimates. Likewise, for six out of the seven additional analyses with plausible evidence of selection bias, the exposures (education, BMI and alcohol) were associated with sex in our negative control analysis.

For the remaining 222 exposure-outcome pairs not reported in Table 3, there was little evidence of bias. This included several pairs where the MR exposure was one of the four variables related to sex in the unweighted GWAS. For those pairs, our negative control MR analysis suggested the existence of bias, but the confidence intervals of the weighted and unweighted GWAS overlapped. It is likely that estimates from these exposure-outcome pairs exhibit a small degree of bias, which would only be noticeable with larger sample sizes.

In this dataset, sex was the only variable available for use as a negative control outcome. Nevertheless, based on our previous analysis, it is worth reflecting on whether sex is a suitable negative control in this context. As discussed previously, biological sex is unlikely to be causally downstream of the 19 traits included in our analyses or their confounders. Sex could plausibly be a cause of some of these traits (e.g. height, alcohol, smoking), and it also may affect confounders, though this is harder to assess given that we used 234 exposure-outcome pairs and each pair will be subject to different confounding effects. On the other hand, the relationship of sex with selection is easier to assess: in the unweighted UK Biobank sample used by Schoeler et al. (2023), 54.4% of participants were women. This proportion was higher than in their HSE sample (50.6%), indicating a potential effect of sex on participation. Taken together, these observations suggest a likely *N*_*Y*_ *→ S* effect, plausible *N*_*Y*_ → *X* and *N*_*Y*_ → *Y* effects and some uncertainty over a *N*_*Y*_ → *U* effect. A negative control outcome with these effects will be similar to variable 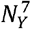 in Table 1. This is not a perfectly valid negative control outcome, as it may mistakenly suggest bias if the genetic instruments associate with selection in UK Biobank in other ways except through their effect on the MR exposure, outcome and confounders (the “*G* → *S*” scenario in Table 1).

To ensure that this does not happen, we repeated our MR and negative control analyses excluding SNPs associated with participation in UK Biobank. We identified such SNPs in two ways: (i) using summary statistics from the GWAS for UK Biobank participation conducted by Schoeler et al. (2023), and (ii) by applying the mtCOJO method (Deng and Pan 2017) to adjust the SNP-participation summary statistics for each exposure and outcome. More details are provided in the Supplement. The results of these sensitivity analyses were similar to those reported in Tables 2 and 3, giving us little reason to be concerned about the validity of sex as a negative control outcome in this application. Note that these sensitivity analyses relied on the availability of a GWAS for UK Biobank participation. When working with other datasets, such a GWAS may not be available, and sensitivity analyses will be harder to implement; we therefore reiterate that caution is needed when using age and sex as negative controls.

Finally, we reiterate that our negative control analysis cannot detect selection bias due to heterogeneity. On the other hand, a comparison of the weighted and unweighted analyses should be able to detect both types of selection bias, with the caveat that the weighting model must be correctly specified. Heterogeneity could be investigated further with access to individual-level data, but this is beyond the scope of our manuscript.

The R code used to conduct this real-data application is available at the GitHub repository: https://github.com/agkatzionis/Negative-controls-in-MR.

## Discussion

Negative controls are a useful diagnostic, both in traditional regression analyses and in MR analyses. Indeed, Davies et al. (2017) suggested using negative controls as one way to compare and triangulate evidence from regression analyses and MR, given that the two methods are subject to different forms of bias. In this manuscript, we have explored what makes a variable a suitable negative control outcome for collider bias in MR studies. Using causal diagrams, we have shown that a negative control outcome should be related to confounders of the MR exposure and outcome. This means that a variable can be a valid negative control regardless of its effect on study participation, as demonstrated in Table 1. We have also explored the use of age and sex as negative control outcomes for MR studies. Finally, we have argued that selection bias due to heterogeneity can also affect MR studies, but it is not possible to detect this form of bias using negative control outcomes.

Several types of negative control analyses for MR studies have been proposed in the literature, including negative control exposures (Yang et al. 2021), negative control instruments (Danieli et al. 2024) and negative control populations (Davies et al. 2017, Dukes et al. 2025). For example, MR analyses using alcohol consumption as an exposure have sometimes used women from East Asian populations as a zero-relevance group, as these women are very unlikely to consume alcohol (Chen et al. 2008, Cho et al. 2015). This can be viewed as a negative control population MR analysis. In this manuscript, we have focused on negative control outcomes, but other types of negative controls may also be useful for detecting selection bias.

In principle, there are two ways to perform a negative control MR analysis: either as an MR analysis between the exposure and the negative control or as a standard regression analysis between the instrument and the negative control (Danieli et al. 2024). Here, we have implicitly assumed that an MR analysis is used; we implemented both approaches in a small-scale simulation used to inform Table 1 and they had a similar performance. In addition, Sanderson et al. (2021) suggested using an MR analysis between the outcome and the negative control to complement the one between the exposure and the negative control when studying population stratification. For selection bias, these two analyses will yield similar results if the MR exposure affects the outcome and the exposure instruments are used for the outcome. Moreover, in this work we have only used negative control outcomes as a diagnostic to detect selection bias. Some authors have gone further and considered the use of negative controls to establish identification and estimate the MR causal effect in the presence of bias (Danieli et al. 2024, Dukes et al. 2025), although this inevitably requires strong assumptions.

Positive control analyses, where the strength of the exposure-positive control effect is known a priori, have also been used in the literature. Although we have not discussed positive controls in this manuscript, we expect that similar criteria are needed to identify a suitable positive control outcome as for a negative control outcome. Sometimes, positive control analyses are implemented when only the direction (and not the magnitude) of the exposure’s effect on the positive control outcome is known; such positive controls are easier to identify but are unable to detect biases that are not strong enough to change the direction of the exposure-positive control effect.

In practice, MR studies have to deal with multiple sources of bias. A valid negative control outcome should therefore be able to detect bias from many sources. In this manuscript we focused on detecting selection bias, but choosing a negative control outcome that is related to confounders of the MR exposure and outcome also helps in detecting other biases, such as population stratification (Sanderson et al. 2021). Other forms of bias, such as pleiotropy, may be harder (though not impossible) to demonstrate using negative controls. It is therefore advisable to combine negative controls with other bias adjustments and sensitivity analyses when performing MR.

Identifying a valid negative control variable in applications can be hard. Davies et al. (2017) proposed the use of outcomes that occurred before individuals were exposed to the exposure of interest; this may not be possible when studying lifelong exposures such as blood pressure or BMI, and even if such outcomes can be identified they may still be subject to bias due to pleiotropy. Some authors have conducted regression analyses using partner data as negative controls when studying the effects of maternal exposures on pregnancy-related and offspring outcomes (Davey Smith 2008, Brew et al. 2017, Cohen et al. 2019). In their exploration of bias due to population stratification, Sanderson et al. (2021) used self-reported tanning ability and natural hair colour as negative control outcomes, since these traits are genetically inherited and likely to be affected by population stratification but otherwise unrelated to common MR exposures. In most cases, however, identifying a valid negative control will be difficult, and as we have shown, will involve careful consideration of the confounding structure between the exposure and outcome of interest. It is perhaps for this reason that some authors have used age and sex as negative control outcomes. Here, we have argued that these variables can allow researchers to detect collider bias in some applications, but are not guaranteed to be valid negative control outcomes and therefore should be used with caution. In the Supplement, we briefly review the use of age and sex as negative control outcomes in the non-linear MR literature and discuss implications of our work. Finally, further challenges regarding the use of age and sex arise when considering other types of bias; for example, it is hard to imagine how these variables could be used as negative controls to study bias due to population stratification.

The advice we have provided in our manuscript will help applied researchers using negative control outcomes and will facilitate further use of negative controls in MR and instrumental variable analyses.

## Data Availability

The real-data analysis conducted as part of this study used summary-level data from the UK Biobank, which are publicly available at: https://drive.google.com/drive/folders/1QMq9Y_BDK-9o0TlaJL-fwaqQl4BbR6Ek
The code used to analyse the data has been made available by the authors at the GitHub repository: https://github.com/agkatzionis/Negative-controls-in-MR?tab=readme-ov-file

https://drive.google.com/drive/folders/1QMq9Y_BDK-9o0TlaJL-fwaqQl4BbR6Ek

