## Supplementary File for "Using Negative Control Outcomes to Detect Selection Bias in Mendelian Randomization Studies"

**Instrumental Variable Assumptions for Conditional Instruments**

In this manuscript, we have used the definition of (Pearl 2009). For completeness, we present Pearl’s (graphical) definition of the IV assumptions in its original form: $G$ is a valid instrument for the effect of exposure $X$ on outcome $Y$ in a directed acyclic graph (DAG) $\mathcal{G}$ if there exists a set of observed variables $C$ such that:

- (IV1): $G$ is conditionally associated with $X$ given $C$ in diagram $\mathcal{G}$.
- (IV2): $G$ is conditionally independent of $Y$ given $C$ in the “manipulated DAG” derived from $\mathcal{G}$ by eliminating the $X-Y$ causal effect.
- (IV3): The set $C$ does not include any variables causally downstream of the outcome $Y$.

If these assumptions are satisfied, $G$ is a valid instrument conditional on the covariates $C$. In the main part of our manuscript, we simplified this by assuming that $C=\emptyset$ and did not consider conditional instruments. Although we condition on the selection indicator $S$ throughout the manuscript, this represents limitations in data availability and not a conscious decision made to render the instrument $G$ valid.

**Different Versions of Instrumental Variable Assumptions and Selection Bias**

As mentioned in our manuscript, there are multiple ways of defining the instrumental variable assumptions. Here, we make the point that not every version of the IV assumptions in the literature is equally well suited to studying selection bias. For example, consider the following version, commonly used in genetic epidemiology (Sanderson et al. 2022):

- (A1): The instrument is (robustly) associated with the exposure.
- (A2): There are no causes of the instrument that also influence the outcome through mechanisms other than the exposure of interest (no confounders of the instrument and the outcome).
- (A3): The instrument does not affect the outcome other than through the exposure and does not affect any other trait that has a downstream effect on the outcome of interest.

The popularity of this set of assumptions in MR lies in their interpretation in terms of common forms of bias: weak instrument bias results in near-violations of assumption (A1), population structure and dynastic effects can violate assumption (A2) and pleiotropy can violate (A3).

Selection bias is a distinct type of bias to confounding, pleiotropy and weak instrument bias and it may be hard to recognise it as a violation of assumptions (A1)-(A3). For example, suppose that we are conducting an MR analysis subject to selection as in the causal diagram of Figure S1a. It is easy to verify that assumptions (A1)-(A3) are satisfied in this diagram: the instrument causes the exposure, there are no confounders of the instrument and the outcome, and the only causal path connecting the instrument to the outcome is via the exposure. On the other hand, assumption IV2 of Pearl (2009) is violated in this diagram, as the path $G\to X\leftarrow U\to Y$ becomes open conditional on $S$.

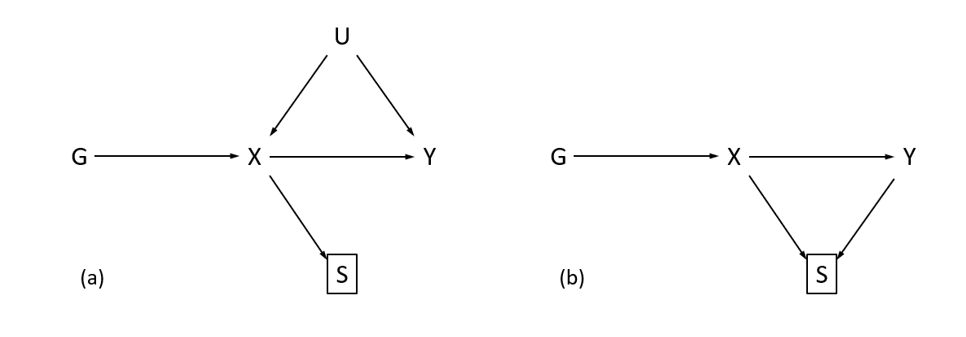

Figure S1: Two causal diagrams in which different versions of the IV assumptions may yield different answers to the question whether the IV assumptions are satisfied.

Another commonly used version of the IV assumptions is:

- (B1): The instrument $G$ is associated with the exposure $X$.
- (B2): The instrument $G$ is independent of any confounders $U$ of the exposure and outcome.
- (B3): The instrument $G$ is conditionally independent of the outcome $Y$ given the exposure $X$ and confounders $U$.

This version can be used to identify most instances of selection bias as violations of the IV assumptions. For example, it is easy to check that this set of assumptions is violated in Figure S1a, as conditioning on $S$ opens the path $G\to X\leftarrow U$ and induces correlation between the instrument and the confounder. However, consider the (admittedly unlikely) MR analysis of Figure S1b, where there is no confounding, but both the exposure and the outcome affect study participation. In this scenario, the instrument-outcome association is biased because the path $G\to X\to S\leftarrow Y$ becomes open in the selected sample. Nevertheless, both assumptions (A1)-(A3) and assumptions (B1)-(B3) appear to be satisfied.

This apparent contradiction is relatively inconsequential. Regardless of how the IV assumptions are defined, the MR analyses of Figure S1 are biased. However, a researcher using Pearl’s assumptions would conclude that this bias constitutes a violation of the IV assumptions while another researcher using assumptions (A1)-(A3) or (B1)-(B3) might conclude that it is a separate source of bias altogether. To the best of our knowledge, this occurs because these assumptions are simplified versions of more formal sets of assumptions defined using counterfactual theory. For example, the counterfactual version of assumptions (A1)-(A3) is presented in Hernan and Robins (2006). These assumptions are violated in both causal diagrams of Figure S1, confirming that formally, collider bias can violate the IV assumptions.

**Heterogeneity and Graphical Criteria for Detecting Selection Bias in Regression Analyses**

As we have argued in the main part of our manuscript, there are two types of selection bias: collider bias and bias due to heterogeneity. When referring to heterogeneity, we allow the term to incorporate a variety of different concepts including effect modification, interaction and non-collapsibility.

Collider bias is easy to represent and hence detect in causal diagrams: the bias occurs because conditioning on one variable induces (or modifies the) associations between its ancestors. On the other hand, bias due to heterogeneity is not a graphical concept and is harder to represent in causal diagrams. Continuing our discussion about the use of different versions of IV assumptions for selection bias, we would argue that the use of graphical sets of assumptions is not enough to investigate heterogeneity. Instead, some applied researchers may find it easier to think of this form of bias as a violation of the modelling assumptions made to estimate the MR causal effect, and not a violation of the IV assumptions. For example, the bias could arise due to the presence of interactions between the exposure and other variables in the outcome model, if these interactions are not accounted for.

Nevertheless, in standard regression analyses, several authors have attempted to develop graphical criteria to detect selection bias of either form. Daniel et al. (2012) proposed a set of generalized backdoor path rules that can be used to detect selection bias when estimating the interventional distribution of an outcome $Y$ given an intervention that fixes the value of an exposure $X$ to $x$. In related work, Bareinboim et al. (2014) derived identification results both for the (observational) conditional distribution $p(Y=y|X=x)$ and for the interventional distribution $p(Y=y|do\left( X=x \right))$. In particular, they showed that the observational distribution can be recovered in the presence of selection bias if $Y\perp S|X$. Building on those results, Mathur et al. (2025) investigated how to construct sufficient adjustment sets for selection bias.

In principle, such results could be used to detect selection bias in MR studies given a particular causal diagram, including both collider bias and bias due to heterogeneity. For example, consider the two DAGs of Figure S2. Application of the standard rules of d-separation in these causal diagrams suggests that collider bias does not occur, as there is only one cause of the conditioned variable $S$ in each diagram. However, selection bias due to effect heterogeneity can still occur if the $X-Y$ causal effect is modified by $U$ (in Figure S2a) or $M$ (in Figure S2b). Accordingly, an application of the results of Bareinboim et al. (2014) suggests that bias can occur in either of the two diagrams, as the outcome is not independent of the selection variable. Note that Figure S2a represents the scenario where $U$ affects selection, which was included in Table 1 of our manuscript and claimed to be free of bias; this is because we were focusing on collider bias in that table.

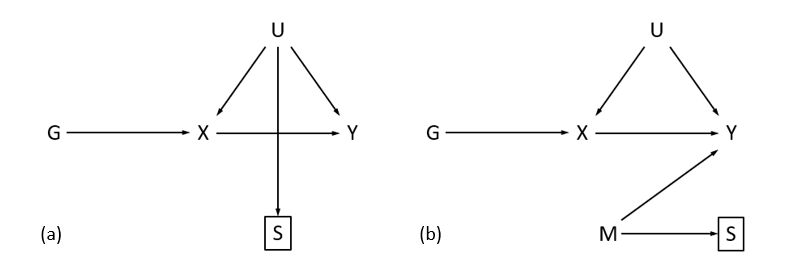

Figure S2: MR analyses where collier bias is not induced by conditioning on selection, but selection bias due to effect modification can still be induced.

It is also worth noting that the graphical criteria discussed above are designed to detect bias in the (conditional or interventional) distribution of the outcome given the exposure. In MR analyses, the parameter of interest is often more specific and represents an average causal effect under homogeneity, or an average causal effect in compliers under monotonicity (Hernan and Robins 2024). Therefore, even in analyses where there is selection bias in the outcome distribution, that bias may not affect the MR estimate itself. Graphical criteria for selection bias in the MR estimand are harder to construct without making parametric modelling assumptions. Bareinboim and Pearl (2011) discussed conditions for estimating exposure-outcome odds ratios in the presence of selection.

The causal diagram of Figure S2b is worth elaborating on, as it represents a selection mechanism not covered in the main part of this manuscript. In Table 1, we considered scenarios where the MR instrument, confounder, exposure and outcome are causes of selection. In practice, however, these variables can relate to selection via common causes, without directly causing it. In this case, collider bias will not be induced: in Figure S2b, conditioning on $S$ does not open any paths from $G$ to $Y$. Hence, we did not consider these selection mechanisms when studying collider bias, although they can still induce selection bias due to heterogeneity.

**Use of Age and Sex in Non-linear MR**

Recently, negative controls have become central to an ongoing debate about the performance of non-linear MR methods. Here, we give a summary of how age and sex have been used in this setting and discuss possible implications of our work. The debate centres around the performance of two non-linear MR methods, namely the residual (Staley and Burgess 2017) and the doubly ranked (Tian et al. 2023) stratification method. Recently, Wade et al. (2023) and Hamilton et al. (2024) used age and sex as negative control outcomes to assess the performance of these methods with real data. Wade et al. (2023) conducted a non-linear MR analysis of the effects of BMI on sex using the residual and doubly ranked methods. Hamilton et al. (2024) conducted four similar analyses, using either BMI or vitamin D levels as the exposure and either age or sex as outcome. The authors observed spurious effects, especially when the exposure was BMI. These findings could be due to violations of the specific assumptions made by the non-linear MR methods (Hamilton et al. 2025), or they could be explained by selection bias and other sources of bias that are inherent in the datasets used.

Both Wade et al. (2023) and Hamilton et al. (2024) used data from the UK Biobank, and it is known that females and older individuals had higher participation rates in the study (Fry et al. 2017). It is therefore possible that these negative control analyses may be biased by selection. A similar argument was made by Schooling and Yang (2024), who noted that the negative control analysis of Wade et al. (2023) may be affected by survival bias. Given that Wade et al. (2023) used data from UK Biobank, we suspect that the bias is more likely to be caused by selective participation than survival, but these two types of bias are structurally similar. Nevertheless, both Wade et al. (2023) and Hamilton et al. (2024) also conducted standard (linear) MR analyses, which ought to be susceptible to selection bias as well; Hamilton et al. (2024) observed no effects of BMI and Vitamin D on age and sex, while Wade et al. (2023) observed an effect of much smaller magnitude than in the non-linear MR analysis. This may suggest that non-linear MR is more susceptible to selection bias than linear MR, and we advocate for further research on this topic.

**Additional Analyses for the Real Data Application**

In our real-data application, we investigated selection bias in pairwise MR analyses between 19 traits in UK Biobank, using sex as a negative control outcome. As we have argued, when using sex as a negative control outcome, it is useful to also assess its relationship with the MR exposure, outcome, confounders and study participation.

Sex is likely to affect UK Biobank participation, given that women make up 54.4% of the UK Biobank sample, compared to approximately 50.6% in the UK population. Given that we are conducting a large number of analyses here, it would be difficult to go through all of them and elaborate on the relationship between sex and the exposure, outcome and confounders in each analysis. Nevertheless, we suspect that sex is likely to relate to at least some of the 19 traits in our UKB analysis (e.g. height, smoking, alcohol consumption). As biological sex is determined at conception and not caused by other variables, these relationships can be depicted as a causal path from sex to these variables. Therefore, sex can be graphically represented as a variable $N_{Y}$ that has a direct effect on selection $S$ and possible effects on $X, U, Y$. Note that if there are multiple causal pathways linking our negative control outcome to the other variables (exposure, outcome, instrument, confounders, selection), the negative control analysis will be biased if at least one of these pathways induces bias. This means that sex will exhibit a similar bias pattern in our applied analyses as the negative control variable $N_{Y}^{7}$ in Table 1 of the manuscript. Therefore, it can provide a correct assessment of selection bias for 15 out of the 16 possible selection mechanisms and only yields misleading results in the “$G\to S$” scenario where the instrument affects selection independently of the exposure, outcome and confounders. Thus, for our real-data analyses, if we can exclude this scenario, it will be easier to justify the use of sex as a negative control outcome.

Schoeler et al. (2023) conducted a GWAS for participation in the UK Biobank, which can be used for this purpose. Summary statistics from this GWAS are available alongside the summary statistics for sex and the 19 traits used in our application. We used these summary statistics for participation to conduct two sensitivity analyses. For our first analysis, we simply identified SNPs associated with participation in the Schoeler et al. (2023) GWAS at a genome-wise significance level of $5\times{10}^{-8}$. There were a total of 2474 SNPs associated with UK Biobank participation, before clumping. We excluded those SNPs from consideration and repeated our MR and negative control analyses using the remaining SNPs. The results of the negative control analyses are reported in Table S4, while the full MR results are given in the Supplementary Data file.

| Exposure | Number of SNPs | Unweighted MR | | | Weighted MR | | |
| --- | --- | --- | --- | --- | --- | --- | --- |
|  |  | Estimate | 95% CI | P value | Estimate | 95% CI | P value |
| BMI | 221 | -0.002 | (-0.005, 0.000) | 0.031 | -0.002 | (-0.004, 0.001) | 0.202 |
| Height | 538 | 0.000 | (0.000, 0.001) | 0.190 | 0.000 | (-0.001, 0.001) | 0.594 |
| Alcohol | 21 | 0.005 | (0.002, 0.007) | 0.001 | 0.001 | (-0.002, 0.004) | 0.553 |
| Smoking | 39 | 0.013 | (-0.039, 0.065) | 0.630 | 0.055 | (0.005, 0.104) | 0.032 |
| Coffee | 19 | -0.001 | (-0.014, 0.013) | 0.902 | -0.008 | (-0.024, 0.008) | 0.346 |
| Fruit | 12 | 0.010 | (-0.016, 0.037) | 0.448 | 0.008 | (-0.040, 0.055) | 0.757 |
| Education | 207 | 0.007 | (0.000, 0.014) | 0.066 | 0.012 | (-0.001, 0.025) | 0.065 |
| LDL | 113 | 0.002 | (-0.007, 0.011) | 0.638 | 0.001 | (-0.013, 0.015) | 0.901 |
| SBP | 118 | -0.001 | (-0.001, 0.000) | 0.125 | -0.001 | (-0.002, 0.000) | 0.105 |
| Non-cancer | 14 | -0.025 | (-0.051, 0.002) | 0.067 | -0.020 | (-0.055, 0.015) | 0.264 |
| Diabetes | 37 | -0.068 | (-0.184, 0.049) | 0.256 | -0.103 | (-0.282, 0.077) | 0.262 |
| Reaction Time | 14 | -0.001 | (-0.001, 0.000) | 0.017 | 0.000 | (-0.001, 0.000) | 0.246 |
| Insomnia | 18 | 0.017 | (-0.061, 0.095) | 0.674 | 0.054 | (-0.053, 0.160) | 0.322 |

Table S4. Estimates of the effect of various traits on sex in the negative control analysis excluding the 2474 SNPs associated with participation at a GWAS level.

Overall, the results of this sensitivity analysis are very similar to those of Table 1 in the main part of the paper: BMI, alcohol and reaction time remained associated with sex in the unweighted GWAS, while the effect of education was attenuated. In fact, the results are identical for 10 of the 13 traits and only differ for height, smoking and education, as these three traits are the only ones for which some of the SNPs originally used as instruments were excluded.

However, this sensitivity analysis has an important limitation: it cannot distinguish between SNPs that directly affect participation (and hence may cause the problematic “$G\to S$” scenario for our negative control analysis) and SNPs that only affect participation via their effects on the MR exposures and outcomes (and therefore discarding them from the MR analyses will provide no benefit, and will instead reduce power). With that in mind, we conducted a second sensitivity analysis using mtCOJO (Deng and Pan 2017). The mtCOJO method can be used to convert marginal summary statistics to conditional ones: given a trait $V_{1}$ of primary interest, an additional set of traits $V_{2},\ldots, V_{K}$ and a set of GWAS summary statistics for each trait, the method can obtain conditional summary statistics for trait $V_{1}$ adjusted for the other traits. Here, we used the method to adjust SNP-participation effects for the effects of the other 19 traits. For each exposure-outcome pair, we first selected the SNPs associated with the exposure at a GWAS threshold in the unweighted GWAS. We then applied mtCOJO to adjust the SNP-participation summary statistics of those SNPs for the exposure and the outcome. We then excluded from further consideration any SNP whose conditional p-value for participation was lower than a Bonferroni-corrected threshold of 0.05 divided by the number of SNPs in the analysis. The remaining SNPs were then used to run the exposure-outcome MR analysis.

The results are reported in Table S5 (negative controls) and in the Supplementary Data file (MR analyses). Again, these largely agreed with our main analysis (Tables 2-3): only a small number of SNPs were discarded and effect estimates were similar. In the negative control analysis, education and reaction time were again observed to affect sex, while for BMI and alcohol the effects attenuated slightly (giving p-values of 0.079 and 0.086 respectively).

| Exposure | Number of SNPs | Unweighted MR | | | Weighted MR | | |
| --- | --- | --- | --- | --- | --- | --- | --- |
|  |  | Estimate | 95% CI | P value | Estimate | 95% CI | P value |
| BMI | 206 | -0.002 | (-0.005, 0.000) | 0.079 | -0.002 | (-0.004, 0.001) | 0.202 |
| Height | 518 | 0.000 | (0.000, 0.001) | 0.285 | 0.000 | (-0.001, 0.001) | 0.594 |
| Alcohol | 16 | 0.003 | (0.000, 0.006) | 0.086 | 0.001 | (-0.002, 0.004) | 0.553 |
| Smoking | 38 | 0.011 | (-0.042, 0.064) | 0.697 | 0.055 | (0.005, 0.104) | 0.032 |
| Coffee | 15 | -0.002 | (-0.018, 0.014) | 0.806 | -0.008 | (-0.024, 0.008) | 0.346 |
| Fruit | 10 | 0.011 | (-0.020, 0.041) | 0.497 | 0.008 | (-0.040, 0.055) | 0.757 |
| Education | 209 | 0.010 | (0.003, 0.017) | 0.007 | 0.012 | (-0.001, 0.025) | 0.065 |
| LDL | 107 | 0.003 | (-0.006, 0.012) | 0.505 | 0.001 | (-0.013, 0.015) | 0.901 |
| SBP | 107 | 0.000 | (-0.001, 0.000) | 0.239 | -0.001 | (-0.002, 0.000) | 0.105 |
| Non-cancer | 11 | -0.031 | (-0.063, 0.000) | 0.051 | -0.020 | (-0.055, 0.015) | 0.264 |
| Diabetes | 37 | -0.068 | (-0.184, 0.049) | 0.256 | -0.103 | (-0.282, 0.077) | 0.262 |
| Reaction Time | 13 | -0.001 | (-0.001, 0.000) | 0.039 | 0.000 | (-0.001, 0.000) | 0.246 |
| Insomnia | 17 | 0.021 | (-0.062, 0.103) | 0.622 | 0.054 | (-0.053, 0.160) | 0.322 |

Table S5. Estimates of the effect of various traits on sex in the negative control analysis applying mtCoJo.

Between our two sensitivity analyses, we conclude that the “$G\to S$” scenario is unlikely to have been the main cause of the associations observed in our negative control analysis. These associations are therefore indicative of (selection) bias in the original MR analyses.

We note that our two sensitivity analyses here relied on having access to a GWAS for UK Biobank participation. When working with other datasets, researchers may not have access to such a GWAS and such sensitivity analyses may be harder to implement. We therefore reiterate our warning that age and sex should be used with caution as negative controls.

Finally, we mention that in practice applied MR studies have to contend with multiple sources of bias. Here, the use of the weighted GWAS of Schoeler et al. (2023) as a reference point allowed us to focus on selection bias. Other possible sources of bias in our application include pleiotropy, weak instrument bias, winner’s curse bias and sample overlap. Methods have been developed to deal with such biases; in the Supplementary Data file, we include results from standard pleiotropy-robust MR methods (MR-Egger, the weighted median and the weighted mode), as well as the MR-lap method (Mounier and Kutalik 2023) that can mitigate bias due to weak instruments, winner’s curse and sample overlap.
